# Pre-existing left bundle branch block and clinical outcomes after transcatheter aortic valve replacement

**DOI:** 10.1101/2023.04.06.23288271

**Authors:** Tetsuya Saito, Taku Inohara, Hikaru Tsuruta, Fumiaki Yashima, Hideyuki Shimizu, Keiichi Fukuda, Yohei Ohno, Hidetaka Nishina, Yoshifumi Nakajima, Masaki Izumo, Masahiko Asami, Toru Naganuma, Kazuki Mizutani, Masahiro Yamawaki, Norio Tada, Futoshi Yamanaka, Shinichi Shirai, Masahiko Noguchi, Hiroshi Ueno, Kensuke Takagi, Yusuke Watanabe, Masanori Yamamoto, Kentaro Hayashida, the OCEAN-TAVI investigators

## Abstract

**Background:** There are currently few reports on pre-existing left bundle branch block (LBBB) in patients undergoing transcatheter aortic valve replacement (TAVR). Nor are there any studies comparing patients with new onset LBBB to those with pre-existing LBBB. This study aimed to investigate the association with pre-existing or new-onset LBBB and clinical outcomes after TAVR.

**Methods:** Using data from the Japanese multicenter registry, 5996 patients who underwent TAVR between October 2013 and December 2019 were included. Patients were classified into 3 groups: no LBBB, pre-existing LBBB and new onset LBBB. The 2-year clinical outcomes were compared between 3 groups using Cox proportinal hazard models and propensity score analysis to adjust the differences in baseline characteristics.

**Results:** Of 5996 patients who underwent TAVR, 280 patients (4.6%) had pre-existing LBBB and new onset LBBB occurred in 1658 patients (27.6%). Compared with no LBBB group, multivariable Cox regression analysis showed that pre-existing LBBB was associated with a higher 2-year all-cause (adjusted hazard ratio [aHR]: 1.39; 95% confidence interval [CI]: 1.06-1.82; p =0.015) and cardiovascular mortality (aHR: 1.62; 95% CI: 1.05-2.54; p =0.027), but also with higher all-cause (aHR:1.43, 95% CI:1.07-1.91; p =0.016) and cardiovascular mortality (aHR: 1.84, 95% CI: 1.14-2.98; p =0.012) than new onset LBBB group. Heart failure was the most common cause of cardiovascular death, with more heart failure deaths in the pre-existing LBBB group.

**Conclusions:** Pre-existing LBBB was independently associated with poor clinical outcomes reflecting increased risk of cardiovascular mortality after TAVR. Patients with pre-existing LBBB should be carefully monitored.

**What is Known?:**

- There are currently few reports on pre-existing left bundle branch block (LBBB) in patients undergoing transcatheter aortic valve replacement (TAVR). Nor are there any studies comparing patients with new onset LBBB to those with pre-existing LBBB.

**What the Study Add?:**

- Patients with pre-existing LBBB not only had a higher mortality than those without LBBB, but also had a worse prognosis than those with new onset LBBB.
- This was because patients with pre-existing LBBB had more heart failure deaths.
- Patients with pre-existing LBBB should be carefully monitored after TAVR. Further investigation will be required to corroborate our findings.

## Introduction

Transcatheter Aortic Valve Replacement (TAVR) is an established therapy in symptomatic severe aortic stenosis (AS) (1). To stratify the risk of patients undergoing TAVR before the procedure, the importance of pre-procedural electrocardiogram is emphasized. Preexisting right bundle branch block (RBBB) is well recognized as a risk factor for the permanent pacemaker implantation (PPI) after the procedure and as an increased risk of all-cause and cardiovascular mortality (2,3). However, the prognostic impact of pre-existing left bundle branch block (LBBB) has not been well investigated.

Several studies investigated the prognostic impact of new-onset LBBB and showed inconsistent results (4–6). The caveat is that these reports exclude patients with pre-existing LBBB and only a few studies were conducted to investigate the impact of pre-existing LBBB on clinical outcomes. Fischer et al. reported pre-existing LBBB was a risk for early PPI after TAVR, but not for late PPI, and had no significant impact on mortality after TAVR (7). Given that the prognostic impact could be different between pre-existing and new-onset LBBB, they should be separately treated in the analysis. Therefore, in the present study, we aimed to investigate the association with pre-existing or new-onset LBBB and clinical outcomes in patients who underwent TAVR for severe AS.

## Methods

### Data source

We analyzed the data from the Optimized transCathEter vAlvular iNtervention (OCEAN) - TAVI registry. A total of 7393 patients were enrolled in the OCEAN -TAVI registry between October 2013 and December 2019. The OCEAN-TAVI registry is a prospective, multicenter, observational registry of patients who underwent TAVR at 20 centers in Japan. The OCEAN-TAVI registry was registered with the University Hospital Medical Information Network Clinical Trial Registry and accepted by the International Committee of Medical Journal Editors (UMIN-ID: 000020423). All study participants provided informed consent, and the registry was approved by the ethics committees of all participating institutions. Patients were followed annually at the participating institutions. The events were site-reported from the participating institutions. For ensuring the consistency, the database was regularly audited by the data committee members. The data underlying this article will be shared on reasonable request.

### Study population

The study flow is presented in Figure 1. We excluded patients who had missing data for electrocardiographic records at either baseline or discharge (n =360), patients with complete RBBB (n =839), patients with unknown native QRS due to ventricular pacing of permanent pacemaker (n =225). As a result, 5996 patients were included in the analyses.

**Figure 1.**
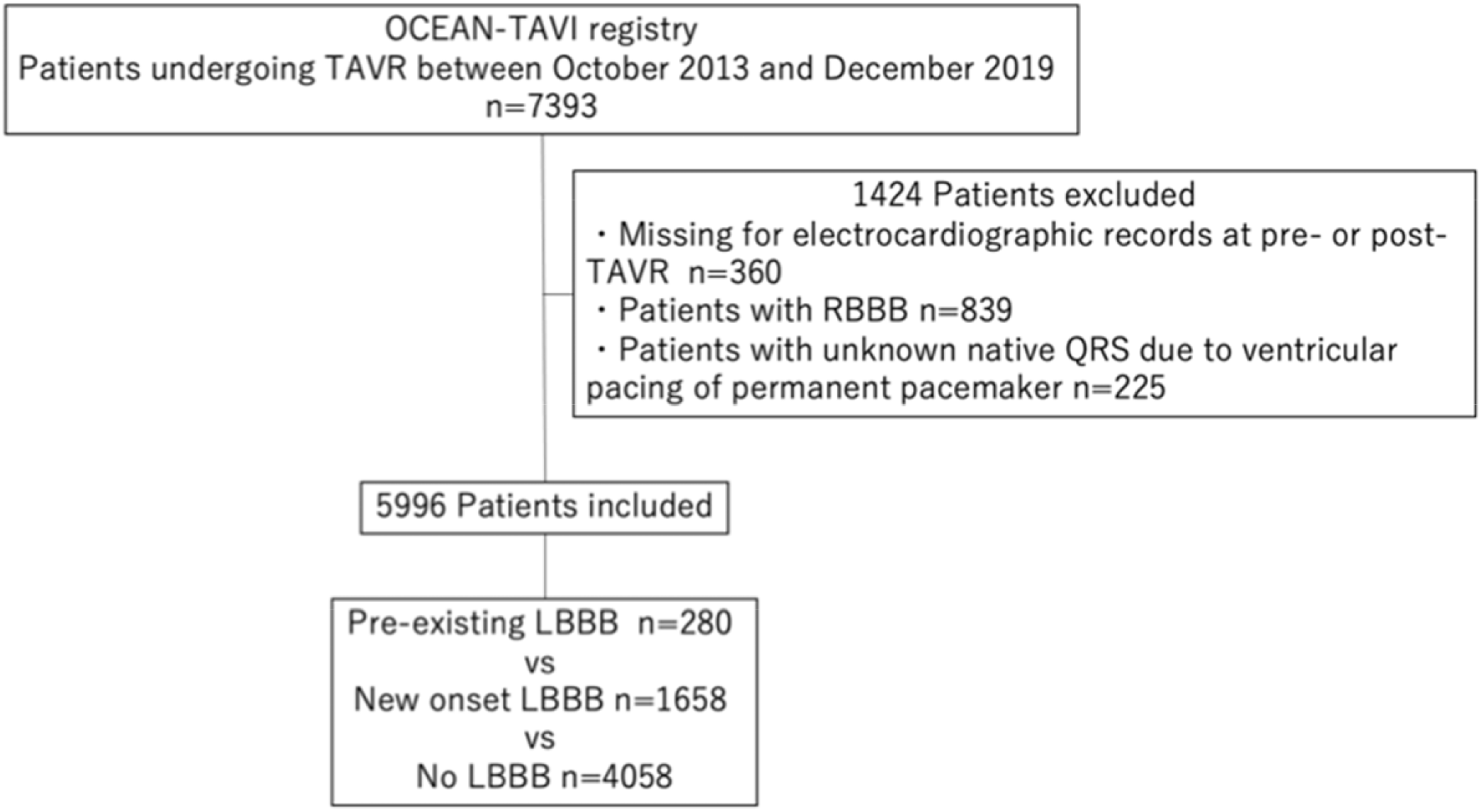
Study flowchart. LBBB indicates right bundle branch block; OCEAN, optimized transcatheter aortic valve implantation; RBBB, left bundle branch block; TAVR, transcatheter aortic valve replacement; and TAVI, transcatheter aortic valve implantation.

### Outcomes

The primary outcomes were 2-year all-cause, cardiovascular and non-cardiovascular mortality after TAVR. The secondary outcomes were death from heart failure, sudden cardiac death (SCD) after TAVR, 30-day mortality and in-hospital complications. All-cause mortality, cardiovascular mortality, complications were defined based on the VARC (the Valve Academic Research Consortium) -2 criteria (8). SCD was defined as a death occurring within one hour of symptom onset if witnessed, or within the past 24 hours if not witnessed, according to the World Health Organization definition. Patients with confirmed sudden death due to terminal disease or non-cardiac causes were not considered SCD (9).

### Electrocardiography and echocardiography

Twelve-lead electrocardiography and transthoracic echocardiography was performed at baseline, before hospital discharge, and at the annual follow-up. The diagnosis of intraventricular conduction disturbances was based on the American Heart Association/American College of Cardiology Foundation/Heart Rhythm Society recommendations for the standardization and interpretation of the electrocardiogram (10). As in previous reports (2,3,7), incomplete RBBB and LBBB were considered normal. All transthoracic echocardiographic parameters were measured according to American Society of Echocardiography guidelines (11-13).

### Statistical analysis

At first, baseline and 30-day outcomes, complications were compared between the three groups: pre-existing LBBB, new onset LBBB and no LBBB (Table 1). Continuous variables were presented as medians and interquartile ranges (IQRs, 25-75%) and compared by analysis of variance (ANOVA) test. Categorical variables were presented as numerical values and percentages and compared by the Pearson chi-square test or the Fisher exact test.

**Table 1.** Baseline characteristics and 30-day outcomes

|  | Pre-existing LBBB<br>n=280 | New onset LBBB<br>N=1658 | No LBBB<br>n=4058 | P value |
| --- | --- | --- | --- | --- |
| Clinical data |  |  |  |  |
| Age, yrs | 85 (82-88) | 85 (81-88) | 84 (81-87) | 0.0018 |
| Male | 88 (31.4) | 374 (22.6) | 1351 (33.3) | <0.001 |
| Body mass index, kg/m<sup>2</sup> | 21.5 (19.1-24.0) | 22.2 (19.6-24.8) | 22.1 (19.6-24.4) | 0.072 |
| Body surface area, m<sup>2</sup> | 1.40 (1.30-1.53) | 1.40 (1.30-1.50) | 1.41 (1.30-1.58) | <0.001 |
| NYHA 3 or 4 | 154 (55.0) | 615 (37.1) | 1658 (40.9) | <0.001 |
| Hypertension | 238 (85.0) | 1405 (84.7) | 3371 (83.1) | 0.24 |
| Dyslipidemia | 150 (53.6) | 941 (56.8) | 2245 (55.5) | 0.47 |
| Diabetes mellitus | 80 (28.6) | 451 (27.2) | 1089 (26.8) | 0.8 |
| Chronic kidney disease | 212 (75.7) | 1162 (70.1) | 2774 (68.4) | 0.023 |
| Previous stroke | 29 (10.4) | 192 (11.6) | 449 (11.1) | 0.77 |
| COPD | 30 (10.7) | 149 (9.0) | 390 (9.9) | 0.59 |
| Peripheral artery disease | 28 (10.0) | 160 (9.7) | 470 (11.6) | 0.092 |
| Coronary artery disease | 95 (33.9) | 505 (30.5) | 1352 (33.3) | 0.099 |
| Previous CABG | 17 (6.1) | 64 (3.9) | 185 (4.6) | 0.2 |
| Atrial fibrillation | 57 (20.4) | 322 (19.4) | 856 (21.1) | 0.36 |
| Permanent pacemaker | 25 (8.9) | 23 (1.4) | 130 (3.2) | <0.001 |
| Clinical frailty scale | 4 (3-5) | 4 (3-4) | 4 (3-4) | 0.096 |
| Beta blockers | 102 (36.4) | 555 (33.5) | 1333 (32.8) | 0.45 |
| RAS inhibitors | 145 (51.8) | 899 (54.2) | 2089 (51.5) | 0.17 |
| STS risk score, % | 7.20 (4.94-11.6) | 6.04 (4.21-8.84) | 6.14 (4.18-9.20) | <0.001 |
| Non-transfemoral approach | 25 (8.9) | 111 (6.7) | 433 (10.7) | <0.001 |
| Local anesthesia | 101 (36.1) | 628 (37.9) | 1468 (36.2) | 0.47 |
| Hemoglobin, g/dl | 11.3 (10.4-12.4) | 11.3 (10.2-12.5) | 11.4 (10.3-12.5) | 0.67 |
| eGFR, ml/min/1.73m<sup>2</sup> | 48.0 (36.9-59.1) | 50.6 (38.5-62.7) | 51.0 (38.5-64.0) | 0.076 |
| Albumin < 3.5 g/dl | 75 (27.1) | 395 (23.9) | 976 (24.2) | 0.51 |
| BNP, pg/ml | 380 (175-782) | 209 (93-455) | 236 (102-526) | <0.001 |
| BNP ≥ 400 pg/ml or NT-Pro BNP ≥ 1600 pg/ml | 148 (54.4) | 556 (34.0) | 1532 (38.3) | <0.001 |
| Echocardiographic data |  |  |  |  |
| Aortic valve area, cm<sup>2</sup> | 0.61 (0.49-0.72) | 0.63 (0.51-0.77) | 0.63 (0.50-0.75) | 0.022 |
| Peak velocity, m/s | 4.30 (3.70-4.80) | 4.50 (4.04-5.08) | 4.50 (4.06-5.10) | <0.001 |
| Mean pressure gradient, mmHg | 42.0 (31.3-54.0) | 47.0 (36.9-61.0) | 47.0 (37.5-61.0) | <0.001 |
| LV end-diastolic dimension, mm | 46.0 (41.0-51.0) | 42.4 (39.0-46.6) | 43.8 (40.0-48.0) | <0.001 |
| LV end-systolic dimension, mm | 32.0 (27.5-40.3) | 27.0 (24.0-31.0) | 28.0 (24.0-33.0) | <0.001 |
| LVEF, % | 51.0 (38.8-61.0) | 64.0 (56.7-68.9) | 62.7 (53.7-68.0) | <0.001 |
| Aortic regurgitation ≥moderate | 35 (12.5) | 144 (8.7) | 442 (10.9) | 0.022 |
| Mitral regurgitation ≥moderate | 49 (17.5) | 156 (9.4) | 465 (11.5) | <0.001 |
| Tricuspid regurgitation ≥moderate | 30 (10.7) | 140 (8.4) | 329 (8.1) | 0.31 |
| 30-day outcomes and complications |  |  |  |  |
| 30-day mortality | 4 (1.4) | 23 (1.4) | 49 (1.2) | 0.83 |
| Stroke | 6 (2.1) | 33 (2.0) | 105 (2.6) | 0.39 |
| Myocardial infarction | 3 (1.1) | 5 (0.3) | 26 (0.6) | 0.15 |
| Vascular complications | 25 (8.9) | 112 (6.8) | 308 (7.6) | 0.33 |
| AKI | 29 (10.4) | 123 (7.4) | 331 (8.2) | 0.22 |
| Bleeding | 35 (12.5) | 198 (11.9) | 592 (14.6) | 0.025 |
| New pacemaker | 8 (2.9) | 92 (5.5) | 124 (3.1) | <0.001 |
| Indexed EOA, cm<sup>2</sup>/m<sup>2</sup> | 1.12 (0.94-1.33) | 1.15 (0.97-1.38) | 1.14 (0.96-1.35) | 0.039 |
| THV mean pressure gradient, mmHg | 10.0 (7.9-13.9) | 10.0 (7.4-13.1) | 10.5 (8.0-14.0) | 0.0039 |
| PVL ≥ moderate | 7 (2.5) | 34 (2.1) | 77 (1.9) | 0.74 |

Data are shown as median (25th-75th percentile) for continuous variables and number (percentage) for categorical variables. AKI indicates acute kidney injury; BNP, brain natriuretic peptide; CABG, coronary artery bypass grafting; COPD, chronic obstructive pulmonary disease; eGFR, estimated glomerular filtration rate; EOA, effective orifice area; LBBB, left bundle branch block; LVEF, left ventricular ejection fraction; NYHA, New York Heart Association; PVL, paravalvular leak; STS, Society of Thoracic Surgeons; and THV, transcatheter heart valve.

The cumulative incidences of all-cause, cardiovascular, and non-cardiovascular mortality, death from heart failure, and SCD were calculated using Kaplan-Meier method. Log-rank test and Cox regression analyses were performed. Univariable Cox regression analyses were performed for 2-year clinical outcomes. Thereafter, multivariable analyses were performed to examine variables that were independently associated with all-cause, cardiovascular, and non-cardiovascular mortality, death from heart failure, SCD. In multivariable analysis, variables associated with mortality based on previous studies were included.

To ensure robust of the results, propensity score analysis was performed. The propensity score was calculated using multinomial logistic model to estimate the probability of pre-existing LBBB, new onset LBBB and no LBBB. The covariables included in the multinomial logistic model are listed in Table S1. Because of the difficulty of propensity score matching for the three groups, inverse probability treatment weighting (IPTW) method was used to analyze 2-year outcomes among the three groups. We used truncated weights at the fifth and 95th centiles to exclude the influence of extreme weights in the weighted cohort. Balance between three groups was assessed by maximum absolute standardized mean difference (SMD).

The loss of cases due to missing values in the multivariable analyses and propensity score analysis was 2.3%. Because the proportion of missing values was small, the multiple imputation was not performed. All statistical analysis was performed using R software version 3.6.1. All tests were 2-sided and p values <0.05 were considered statistically significant.

## Results

### Baseline characteristics and 30-day outcomes

The baseline characteristics and 30-day outcomes of the study are shown in Table 1. Among 5996 patients, baseline electrocardiography showed LBBB in 280 patients (4.6%). Patients with pre-existing LBBB were more symptomatic and had worse renal function, higher BNP, larger left ventricular diameter, lower left ventricular ejection fraction (LVEF), and more severe MR, resulting in higher STS scores. Postprocedural bleeding and new pacemaker implantation were more common in patients with new onset LBBB.

### Mortality

The Kaplan-Meier curves of all-cause, cardiovascular, and non-cardiovascular mortality for the three groups of pre-existing LBBB, new onset LBBB, and no LBBB are shown in Figure 2. The median follow-up period was 686 [IQR:372-744] days. During the follow-up period, 988 patients died of all-cause, 321 of cardiovascular and 667 of non-cardiovascular causes. There was a significant difference among the three groups in 2-year all-cause (log-rank p =0.003) and cardiovascular mortality (log-rank p =0.007). However, there was no significant difference in non-cardiovascular mortality between the three groups (log-rank p=0.13). In multivariable Cox analysis, patients with pre-existing LBBB had higher all-cause (adjusted hazard ratio [aHR]: 1.39; 95% confidence interval [CI]: 1.06-1.82; p =0.015) and cardiovascular mortality (aHR: 1.62; 95% CI: 1.05-2.50; p =0.028) than those without LBBB, and even higher all-cause (aHR: 1.43; 95% CI: 1.07-1.91; p =0.016) and cardiovascular mortality (aHR: 1.84; 95% CI: 1.14-2.98; p =0.012) than those with new onset LBBB. New onset LBBB was not associated with all-cause (aHR: 0.97; 95% CI:0.83-1.13; p =0.75) or cardiovascular mortality (aHR: 0.88; 95% CI:0.66-1.17; p =0.39) (Table 2). Multivariate analysis of noncardiac deaths showed no significant differences among the three groups. The full univariable and multivariable model results were shown in Table S2-4.

**Table 2.** Results of primary outcomes analyses. CI indicates confidence interval; HR, hazard ratio; IPTW, inverse probability treatment weighting; and LBBB, left bundle branch block.

|  |  | Pre-existing vs no LBBB |  | New onset vs no LBBB |  | Pre-existing vs new onset LBBB |  |
| --- | --- | --- | --- | --- | --- | --- | --- |
|  |  | HR (95% CI) | P value | HR (95% CI) | P value | HR (95% CI) | P value |
| All-cause mortality | Unadjusted | 1.44 (1.12-1.87) | 0.0049 | 0.89 (0.77-1.03) | 0.13 | 1.61 (1.22-2.12) | <0.001 |
|  | Multivariable Cox regression analysis | 1.39 (1.06-1.82) | 0.015 | 0.97 (0.83-1.13) | 0.75 | 1.43 (1.07-1.91) | 0.016 |
|  | IPTW analysis | 1.43 (1.09-1.89) | 0.011 | 0.93 (0.80-1.10) | 0.44 | 1.53 (1.13-2.06) | 0.0058 |
| Cardiovascular mortality | Unadjusted | 1.72 (1.14-2.60) | 0.01 | 0.84 (0.65-1.09) | 0.19 | 2.04 (1.29-3.21) | 0.0019 |
|  | Multivariable Cox regression analysis | 1.62 (1.05-2.50) | 0.028 | 0.88 (0.66-1.17) | 0.39 | 1.84 (1.14-2.98) | 0.012 |
|  | IPTW analysis | 1.74 (1.12-2.72) | 0.013 | 0.82 (0.62-1.11) | 0.21 | 2.11 (1.29-3.46) | 0.003 |
| Non-cardiovascular mortality | Unadjusted | 1.31 (0.94-1.81) | 0.11 | 0.91 (0.77-1.09) | 0.34 | 1.46 (1.00-2.01) | 0.05 |
|  | Multivariable Cox regression analysis | 1.26 (0.89-1.78) | 0.19 | 1.01 (0.84-1.21) | 0.89 | 1.24 (0.86-1.79) | 0.24 |
|  | IPTW analysis | 1.19 (0.82-1.72) | 0.34 | 0.97 (0.80-1.17) | 0.79 | 1.22 (0.83-1.81) | 0.31 |

**Figure 2.**
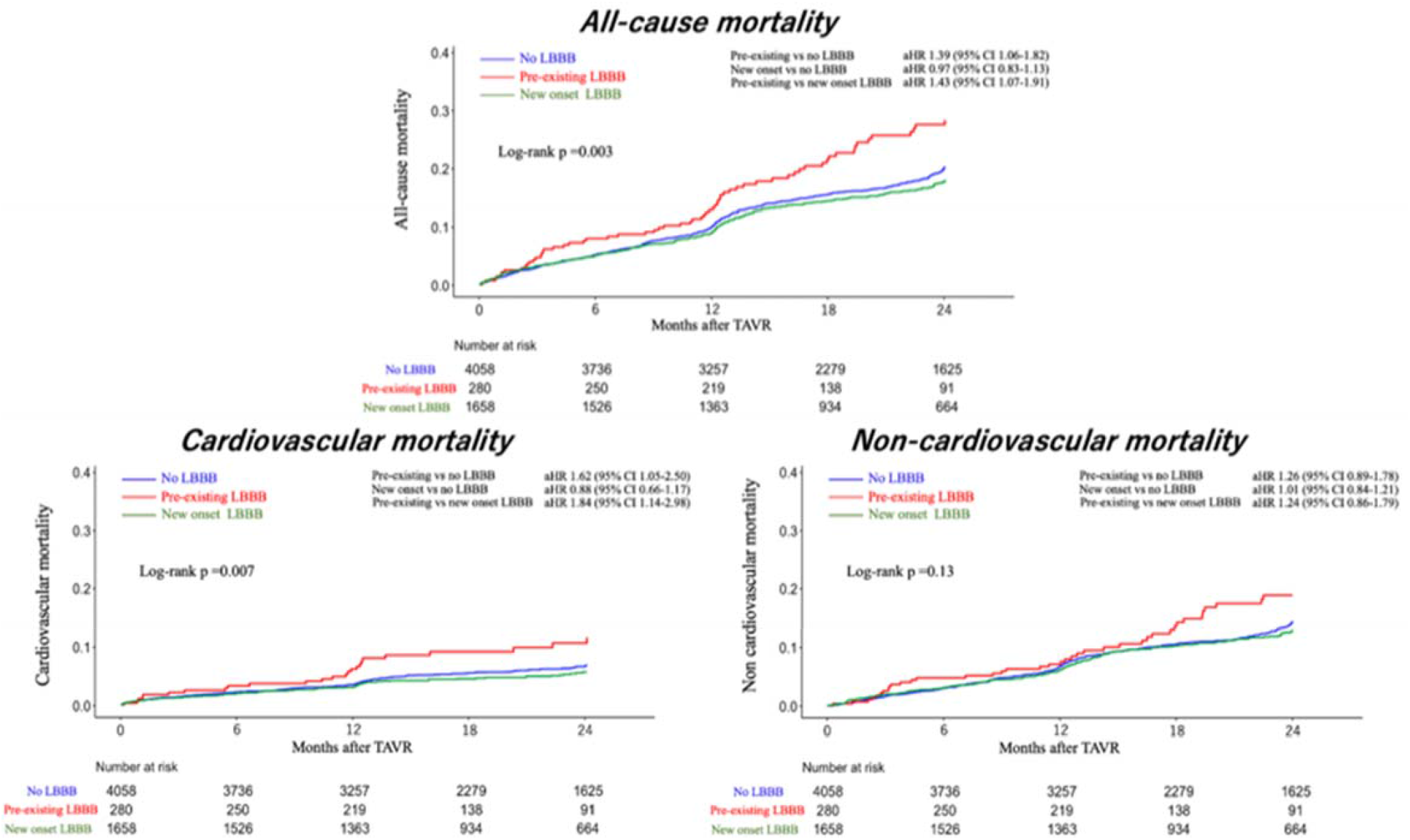
All-cause, cardiovascular and non-cardiovascular mortality in overall cohort. All-cause and cardiovascular mortality of patients with LBBB compared with those without in overall cohort. CI indicates confidence interval; HR, hazard ratio; and LBBB, left bundle branch block.

IPTW analyses were performed. Maximum absolute value of standardized mean difference was less than 0.1 in all examined covariates in the weighted cohort (Table S1). After IPTW, the Kaplan-Meier curves of all-cause, cardiovascular, and non-cardiovascular mortality for the three groups are shown in Figure 3. IPTW analyses showed pre-existing LBBB was associated with higher all-cause (HR: 1.43, 95% CI: 1.09-1.89; p =0.011) and cardiovascular mortality (HR: 1.74, 95% CI: 1.12-2.72; p =0.013) than those without and was associated with higher all-cause (HR: 1.53, 95% CI: 1.13-2.06; p =0.0058) and cardiovascular mortality (HR: 2.11, 95% CI: 1.29-3.46; p =0.003) than those with new onset LBBB. New onset LBBB was not associated with all-cause (HR: 0.93.; 95% CI:0.80-1.10; p =0.44) or cardiovascular mortality (HR: 0.82; 95% CI:0.62-1.11; p =0.21) (Table 2). In the IPTW analysis, there was no significant difference in non-cardiac death between the three groups.

**Figure 3.**
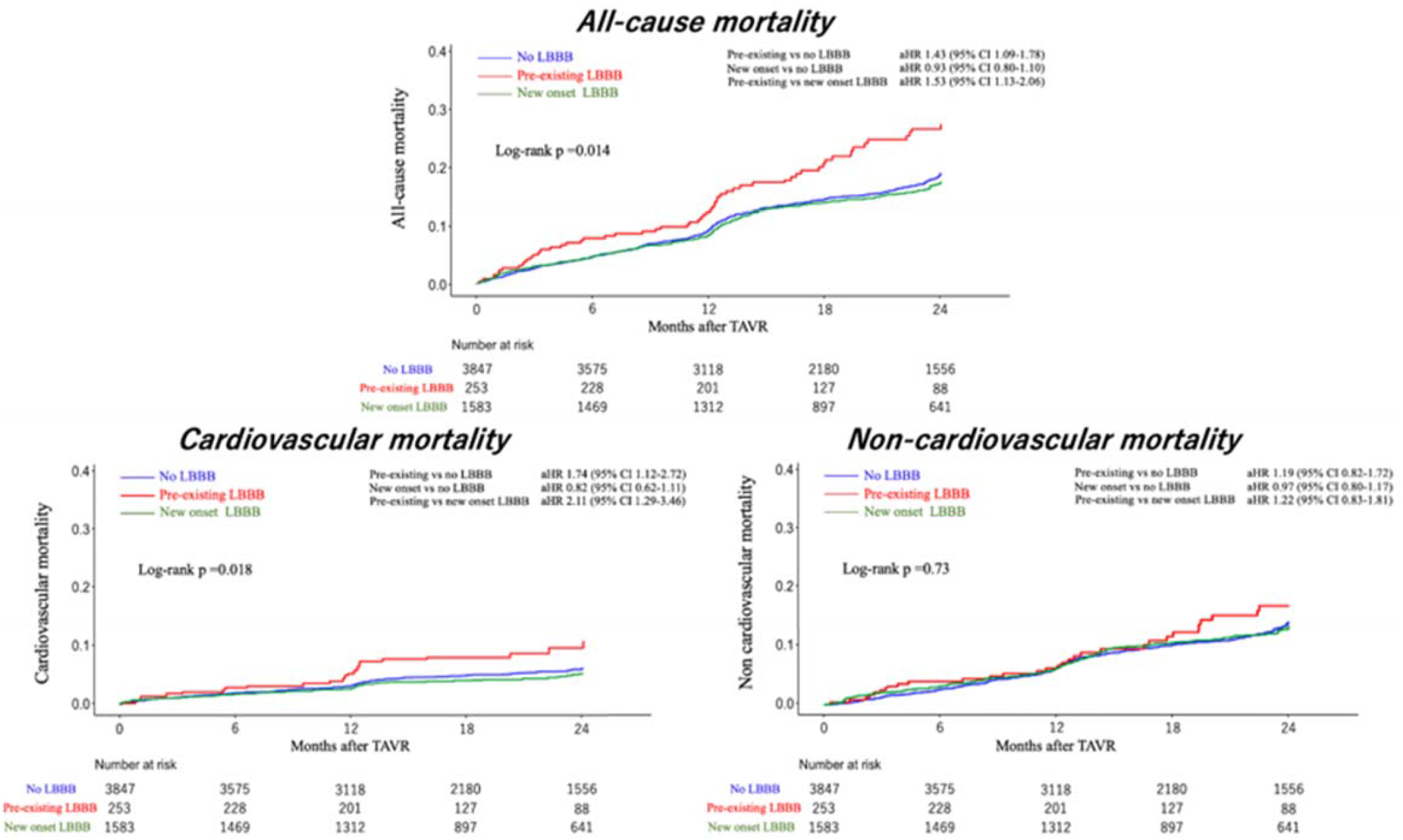
All-cause, cardiovascular and non-cardiovascular mortality in the weighted cohort. All-cause and cardiovascular mortality of patients with pre-existing, new onset, and no LBBB in overall cohort and the weighted cohort. CI indicates confidence interval; HR, hazard ratio; SMD, standardized mean difference, and LBBB, left bundle branch block.

### Death from heart failure and sudden cardiac death

The causes of cardiovascular death in each group are shown in Figure 4. Detailed causes of death are shown in Table S5. Heart failure and SCD were the two leading causes of death in all groups. The Kaplan-Meier curves for heart failure death and SCD before and after IPTW are shown in Figure 5. During the follow-up period, 173 patients died of heart failure, 43 of whom died of sudden cardiac death. Heart failure death was significantly more common in pre-existing LBBB patients (log-rank p <0.001). SCD was not significantly different among the three groups (log-rank p =0.13). In multivariable Cox analysis of heart failure death, patients with pre-existing LBBB was associated with higher risk of heart failure death (aHR: 2.22; 95% CI: 1.34-3.68; p =0.0018) than those without LBBB, and even higher risk of heart failure death (aHR: 2.46; 95% CI: 1.37-4.43; p =0.0025) than those with new onset LBBB. The IPTW analysis likewise showed that pre-existing LBBB not only had more heart failure deaths than no LBBB (aHR: 2.79; 95% CI: 1.68-4.65; p <0.001), but also more heart failure deaths than new onset LBBB (aHR: 3.23; 95% CI: 1.78-5.85; p <0.001). Neither multivariable nor IPTW analysis showed any significant difference in SCD between the three groups (Table 3). The full univariable and multivariable model results were shown in Table S6,7.

**Figure 4.**
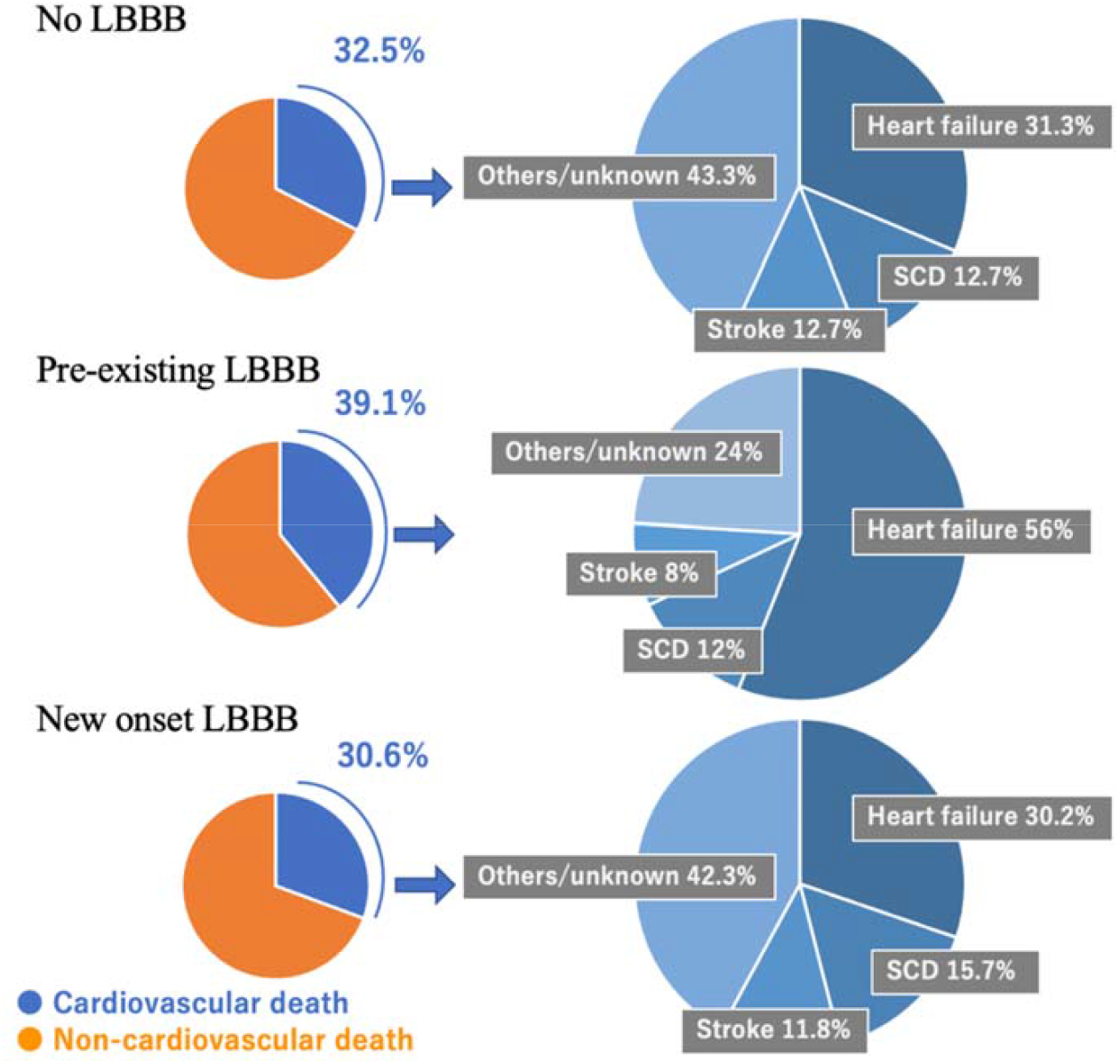
Causes of cardiovascular death. Causes of cardiovascular death in no LBBB, pre-existing LBBB, and new onset LBBB groups. LBBB indicates left bundle branch block, and SCD, sudden cardiac death.

**Figure 5.**
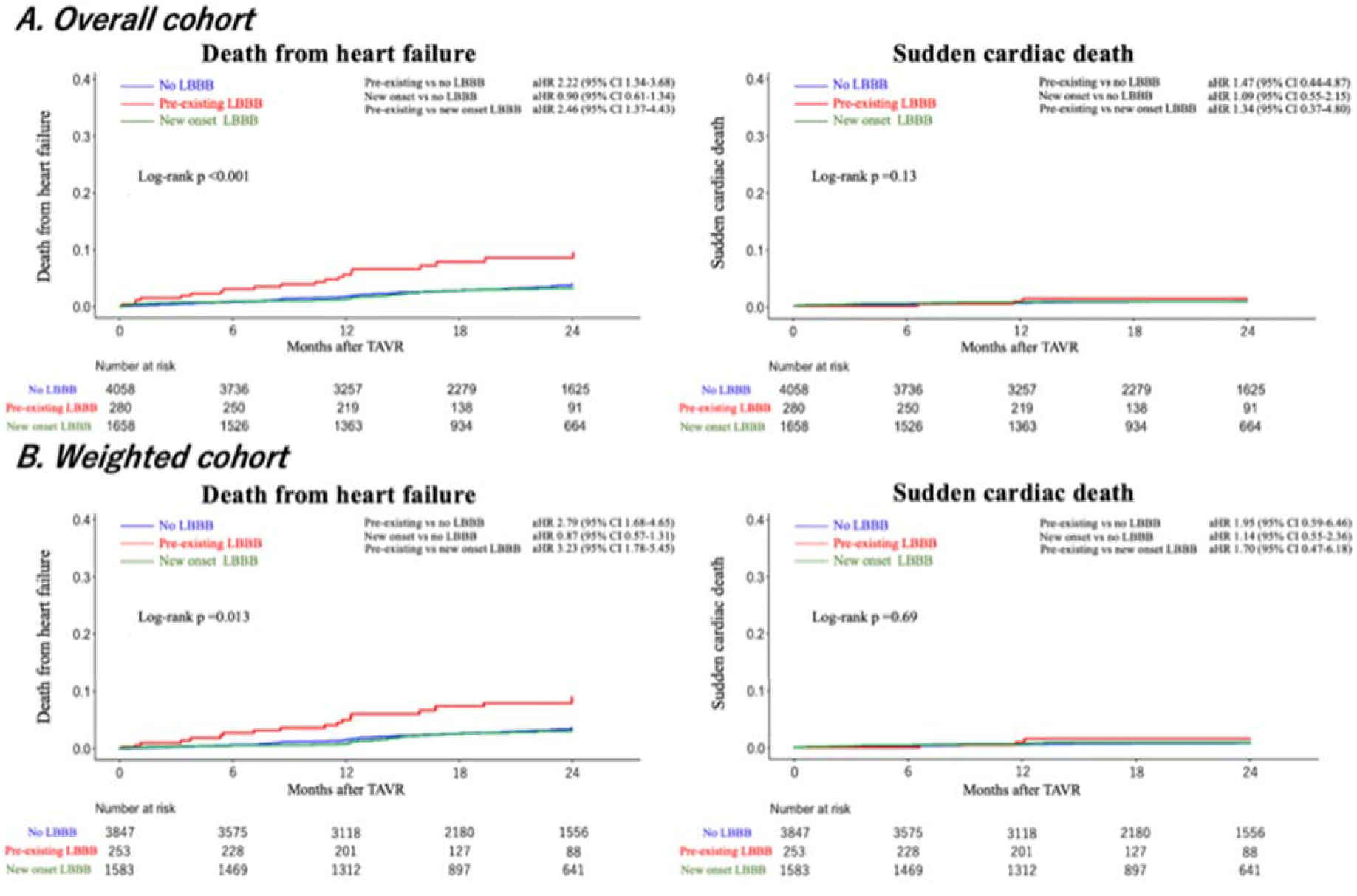
Death from heart failure and sudden cardiac death. Death from heart failure and sudden cardiac death of patients with pre-existing, new onset, and no LBBB in the unweighted cohort and the weighted cohort. CI indicates confidence interval; HR, hazard ratio; and LBBB, left bundle branch block.

**Table 3.**
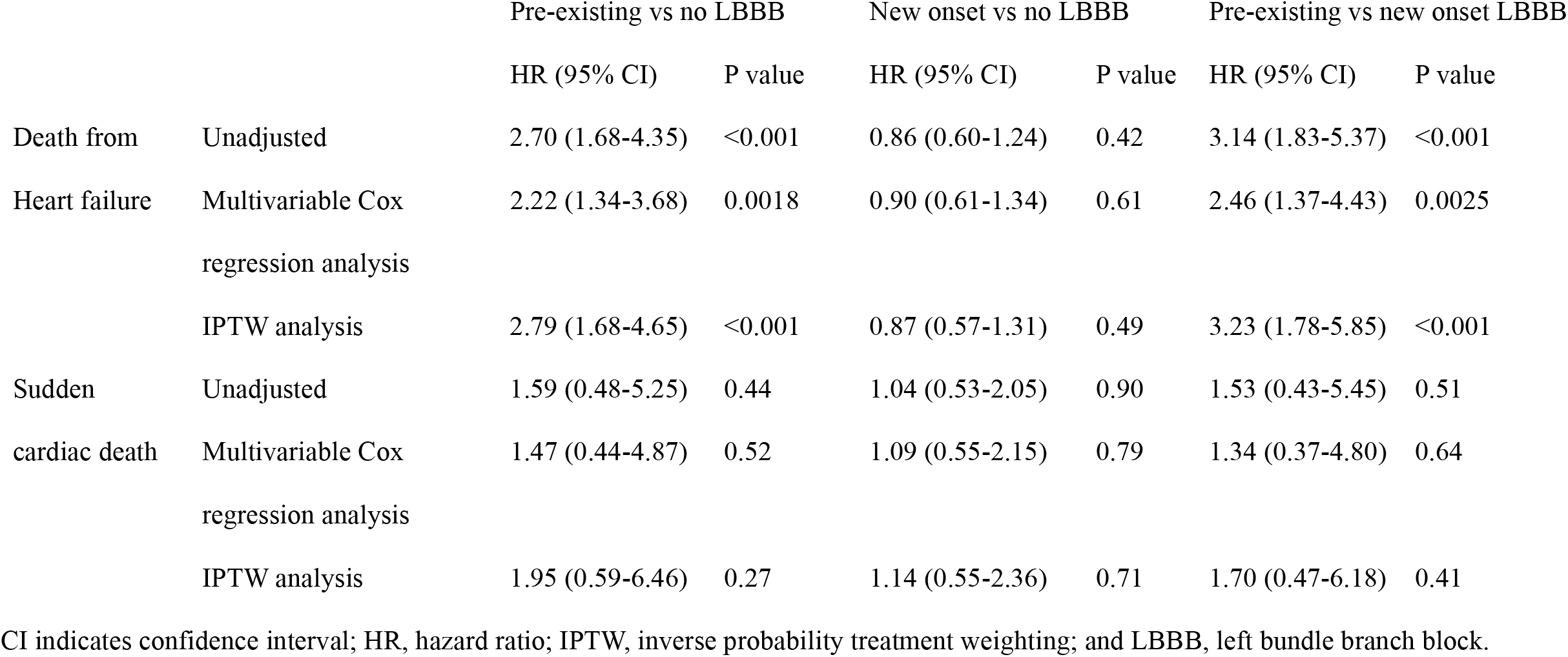
Analyses of death from heart failure and sudden cardiac death

## Discussions

The impact of pre-existing LBBB on patients undergoing TAVR remains unknown. The main findings of our study showed that pre-existing LBBB was associated with poor clinical outcomes after TAVR. Furthermore, patients with pre-existing LBBB had a worse prognosis than those with new-onset LBBB. To the best of our knowledge, the present study is the first to identify the association of pre-existing LBBB and high risk of mortality after TAVR.

Most reports of LBBB in patients who underwent TAVR pertain to postprocedural new-onset LBBB. The prognostic impact of new-onset LBBB is controversial (4-6). However, these reports exclude patients with pre-existing LBBB. As such, there are few studies on the impact of pre-existing LBBB on clinical outcomes after TAVR. Fischer et al. reported pre-existing LBBB is a risk for early PPI after TAVR, but not for late PPI, and furthermore has no significant effect on mortality after TAVR (7). However, in our study, pre-existing LBBB was not a risk for early PPI. In terms of complications, bleeding and pacemaker implantation were more common in patients with new-onset LBBB. However, complications were similar for pre-existing LBBB and no LBBB.

In our study, 4.6% of patients undergoing TAVR had LBBB. This is more than the percentage of LBBB in the general population (14). However, the percentage of pre-existing LBBB patients undergoing TAVR reported so far is around 10% (8), and the incidence in our study is somewhat lower than before. Also, although LBBB increases with age, valvular heart disease and coronary heart disease, cardiomyopathies, myocarditis can be background factors for LBBB. Because patients with severe AS are older and as such are at higher risk than the general population, the rate of LBBB is also likely to be higher than in the general population. In our study, patients with LBBB were more symptomatic and had worse renal function, higher BNP, larger left ventricular diameter, lower LVEF, and more severe MR. LBBB causes dyssynchronous left ventricular activation, left ventricular remodeling, resulting in left ventricular dilatation, low LVEF and MR. Worse outcomes in heart failure patients with LBBB may be expected due to dyssynchronous left ventricular activation. In fact, LBBB is associated with increased all-cause mortality not only in patients with heart failure, but also ischemic heart disease, cardiomyopathy, and even in the general population (14). Based on the findings of prior studies, potential mechanisms of our result may include progression to high-degree AVB, ventricular arrhythmia, or worsening heart failure. However, in our study, many deaths were due to heart failure, not SCD.

In our study, patients with pre-existing LBBB had a worse prognosis than those with new onset LBBB. There have been mixed reports on the possible risk of new onset LBBB with respect to all-cause, cardiovascular death, heart failure hospitalization, and new pacemaker implantation (15). In our study, new onset LBBB was determined only by post-TAVR ECG before or at discharge, and it is possible that some of the patients diagnosed with new onset

LBBB may have improved during follow-up. It has been reported that transient LBBB is not associated with prognosis (16), and our study may have included more transient LBBB than previous studies. In addition, new onset LBBB is caused by mechanical compression of the conduction system by a prosthetic valve, but pre-existing LBBB is due to a failure of one’s own conduction system, which may be hiding cardiomyopathy or other problems that were not fully understood in our study, which may have led to the results. It has been reported that bundle branch block is more common in patients with amyloidosis (17), and since our study excluded RBBB patients, it is possible that there were more patients with amyloidosis in the pre-existing group.

There is no evidence for treatment of LBBB patients after TAVR, whether pre-existing LBBB or new onset LBBB. Further research is needed to determine the timing of device implantation and which devices, including cardiac resynchronization therapy, are best. Since the mortality rate is higher in patients with pre-existing LBBB, it may be important to know whether patients with LBBB after TAVR have pre-existing LBBB or new onset LBBB. Although perioperative risks are not high, patients with pre-existing LBBB should be carefully monitored after TAVR. However, currently, there are no reports other than our study showing similar results. Further investigation is required.

### Study limitations

Our study has several limitations. First, this is a non-randomized, retrospective study using data from a prospective multicenter cohort registry. There is a possibility that unknown and unmeasurable confounding factors exist because this is an observational study. However, we performed sensitivity analyses and only extreme assumptions can distort our results. Second, no detailed electrocardiographic information such as PR interval and QRS duration is available. QRS duration is important, as it has been reported to be associated with prognosis (18). In addition to new onset LBBB, there are also reports that first degree of atrioventricular block is a risk for PPI (19). Third, the indication of PPI during follow up was not collected although the frequency of PPI during follow-up is important. Fourth, there is a lack of procedural and CT data. The length of the membrane septum and the depth of valve implantation are relevant for new onset LBBB and permanent pacemaker implantation (20). Finally, although three-group comparisons were used in this study, multiple comparisons are prone to statistical errors. Therefore, more meticulous studies that consider these should be our future targets.

## Conclusions

Pre-existing LBBB was independently associated with poor outcomes reflecting increased risk of cardiovascular mortality after TAVR. Patients with pre-existing LBBB should be carefully monitored after TAVR. Further investigation will be required to corroborate our findings.

## Data Availability

The data underlying this article will be shared on reasonable request.

## Abbreviations

AS: aortic stenosis
CI: confidence interval
HR: hazard ratio
LBBB: left bundle branch block
TAVR: transcatheter aortic valve replacement

## Acknowledgment

The authors thank all OCEAN-TAVI investigators.

## Source of Funding

The OCEAN-TAVI registry is supported by Edwards Lifesciences, Medtronic, Boston Scientific, Abbott Medical, and Daiichi-Sankyo company.

## Disclosures

Dr. Takagi, Dr. Shimizu are clinical proctors for Edwards Lifesciences. Dr. Asami, Dr. Ohno and Dr. Yashima are clinical proctors for Medtronic. Dr. Ueno, Dr. Tada, Dr. Naganuma, Dr. Mizutani, and Dr. Watanabe are clinical proctors for Edwards Lifesciences and Medtronic. Dr. Hayashida, Dr. Yamamoto, and Dr. Shirai are clinical proctors for Edwards Lifesciences, Abbott Medical, and Medtronic. The remaining authors have nothing to disclose.

## Notes

### Clinical Trial

UMIN-ID: 000020423

### Author Declarations

The OCEAN-TAVI registry was registered with the University Hospital Medical Information Network Clinical Trial Registry and accepted by the International Committee of Medical Journal Editors (UMIN-ID: 000020423). All study participants provided informed consent, and the registry was approved by the ethics committees of all participating institutions. Patients were followed annually at the participating institutions.

